# Predictive Modeling of Clinical Trial Outcomes for Novel Drugs using Digital Twin Patient Cohorts and GenerativeAI

**DOI:** 10.1101/2023.09.11.23295380

**Authors:** Dmitrii K Chebanov, Vsevolod A Misyurin

## Abstract

There is a problem of clinical trial failure, as each new drug should surpass the effectiveness of existing treatment regimens, which becomes increasingly challenging over time. Another significant issue is treating patients who have developed resistance to the current therapies.

Essentially, the use of drug combinations or off-label drug use, where the indication does not match the diagnosis, is akin to an experiment, as there is insufficient data on which drug or combination to use.

This work proposes an approach utilizing computer modeling of patients using gene expression and clinical data. Deep learning and generative adversarial networks are employed as modeling tools. The training data for the algorithms were sourced from publicly available databases such as TCGA and Drugbank.

The modeling is based on the hypothesis of similarity between patients, similarity between drugs, as well as the similarity between individual organs and patient tissues with cell lines, with similarity being computed mathematically. As a result, a patient model is created, where the input consists of drugs and their combinations, and the output provides survival probability values. These model data can be generated in any required quantity with generative adversarial networks (GAN) technology to create observation and control groups. Consequently, it becomes possible to simulate clinical trials, forecasting their outcomes, and, most importantly, optimizing the trial parameters to maximize the likelihood of success.

## Introduction

Many studies have focused on determining whether a patient will surpass the threshold of a defined OS or PFI, leading to classification tasks. Researchers attempt to classify whether a patient will have a positive or negative response to a given therapy [1].

Some approaches predict the response to specific therapeutic agents, but they are limited to a small number of chemotherapy drugs, typically less than ten. Models for other drugs have not been trained [2].

The existing methods for predicting the effectiveness of clinical trials or clinical outcomes are based on different approaches. However, there is limited research available (less than 45 out of 1900) that incorporates transcriptomic analysis data.

Turki proposed the use of a technique known as transfer learning. The approaches utilized included correlation alignment for unsupervised domain adaptation (CORAL), specifically CORAL-SVM, as well as boosting for transfer learning. These methods enabled the extraction of gene expression data most relevant to drug sensitivity. Various strategies were employed by the authors to standardize data across training and testing datasets. Recognizing the limitations of these approaches, the authors developed a novel method based on support vector machines (SVM), xgboost, DeepBoost, and decision trees. This new approach demonstrated enhanced effectiveness, albeit restricted to assessing potential drug sensitivity in patients. The task of predicting patients’ survival time was not addressed [3].

In their intriguing study, López-García employed a densely-connected multi-layer feed-forward neural network (MLNN) model for the prediction of lung-cancer patient’s progression-free interval (PFI). The training dataset consisted of gene expression profiles from patients with various tumor conditions, along with information on the time of occurrence of adverse events. To optimize the MLNN model’s performance, a Bayesian optimization procedure with 100 iterations was applied to collectively fine-tune all hyper-parameters. The trained neural network was then utilized to forecast the PFI duration in lung-cancer patients. Notably, the analysis excluded any influence of medications on patient survival outcomes.

Fascinating research is being conducted regarding the prediction of prostate cancer recurrence [4]. Among several methods applied, the best was random forest model with accuracy 74.2%. Common tasks include tumor classification based on gene expression, although without considering them in conjunction with the administered treatments [5].

It is interesting to provide examples of research where gene expression information was not utilized as input data. For instance, in the study conducted by Motwani, machine learning techniques were employed to predict 5-year all-cause mortality (ACM) in patients undergoing coronary computed tomographic angiography (CCTA) [6]. The performance of this prediction was compared to existing clinical or CCTA metrics. Notably, a smaller set of input features was used for this prediction compared to our study. Specifically, twenty-five clinical and 44 CCTA parameters were assessed. The predictive classifiers for ACM were developed using an ensemble classification approach, specifically employing an iterative LogitBoost algorithm utilizing decision stumps (single-node decision trees) for each feature-selected variable as base classifiers. This method demonstrated greater efficacy compared to classical predictive approaches, achieving an AUC of 0.79 (0.77-0.81).

In the research by Susai et al., vector machine learning techniques were applied to construct predictive models for functional outcomes at the 12-month mark. The input data in this case encompassed cytokine level determinations. It is conceivable that the authors encountered challenges in predicting outcomes within a timeframe of less than 12 months. To explore the relationship between two factors observed over a 6-month period, linear regression models were developed [7].

In another study, Chen et al. employed boosted decision tree, support vector machine, nonparametric random forest, and neural network models to forecast 5-year survivorship in Ewing sarcoma patients [8]. For both cancer-specific survival and overall survival predictions, performance metrics slightly favored the random forest method over the other models, with sensitivities of 77% and 83%, and specificities of 91% and 94%, respectively. While the approach appears intriguing, it is important to note that gene expression data were not utilized. A noteworthy precedent exists in predicting survival within a 1-year timeframe; however, this previous approach did not leverage transcriptomic analysis data. The experimental dataset comprised 14,885 records featuring 22 continuous variables and 43 categorical variables [9]. One of the studies presents a highly efficient approach for predicting the successful completion of Phase 2 clinical trials [10]. However, this approach lacks patient-level detail, including the omission of omics data from the patient profile during training. As a result, it is unable to derive biomarkers that significantly influence outcomes and could serve as criteria for inclusion or exclusion. Consequently, this method is unsuitable for assessing a patient’s relevance to a clinical trial. Furthermore, this approach cannot be employed for quantitatively forecasting the effectiveness of the tested compound in surpassing existing treatment standards.

In this work, we aim to predict the period of event-free survival in terms of the number of months, solving a regression task for each interval type: OS and PFI. Moreover, we will train the model not only for chemotherapy agents but also for targeted drugs, such as small molecule inhibitors, antagonists, and their combinations.

As a result, this model will allow input parameters of patient groups with a specific diagnosis, loading their genomic profiles and clinical data from databases (or generating them), as well as the drugs planned for testing on these patients. The model will predict survival curves for patient groups and the percentages of response categories. The drugs can be any, including those that have never been used for a given diagnosis.

The prediction of overall survival (OS) and disease-free survival (PFI) periods is influenced by both gene expression data, which provide evidence of pathway activation, and clinical data such as patient age and disease stage.

The underlying hypothesis of this study is that patients who are similar in terms of gene expression profiles and clinical data will respond similarly to drugs that share similarities. Therefore, the challenge is to represent these input data in a machine-readable format to mathematically determine the measure of similarity between them.

## Methods

We obtained patient data and drug information from the TCGA (The Cancer Genome Atlas) database located at https://gdc.cancer.gov [11]. Biologicals were excluded, and only small molecules, including targeted therapies and chemotherapeutic agents, were retained. The dataset consisted of information on 3225 patients and the effects of 161 drugs.

According to our hypothesis, the patient’s profile includes three groups of factors, and each of these groups contributes approximately equally to the duration of overall survival (OS) and disease-free survival (PFI) intervals. The first group comprises clinical data such as age, diagnosis, and disease stage. The second group consists of genetic data, specifically gene expression profiles, which capture the functional characteristics of genes. The third group represents the drug or combination of drugs used for patient treatment. Therefore, it is necessary to ensure that all three groups of factors have comparable dimensions of influence.

The following clinical data were available: age, disease stage, diagnosis, gender, duration of OS and PFI intervals, along with information on the occurrence of events. One-hot encoding was employed to encode the clinical variables. For instance, age was divided into eight intervals, and a separate column was created for each interval (‘10-20’, ‘21-30’, …, ‘81-90’). If a patient’s age fell within the first interval, a value of 1 was assigned to the corresponding column, while the rest were assigned 0, and so on. The same approach was applied to disease stages (‘Stage I’, …, ‘Stage IV’), where stages IA and IC were denoted by a value of 1 in the ‘Stage I’ column. Similarly, diagnoses were represented using 33 additional columns with their respective names. Thus, together with binary gender representation, we had a total of 46 clinical feature columns.

For drug data, we obtained information from the DrugBank database [12]. We represented the drug molecules using SMILES (Simplified Molecular Input Line Entry System) notation, which were then converted into 100-dimensional vectors using embedding techniques derived from natural language processing technologies. The RDKit [13], mol2vec [14], and word2vec [15] libraries for Python were utilized for this purpose. To represent combinations of drugs, we employed vector addition of the individual drug vectors. As the outcome measure for prediction, we considered numerical values representing the durations of OS and PFI intervals for each respective task.

The challenge of accurate training lies in the fact that the algorithm cannot distinguish between patients with a short survival period and those who experienced a short follow-up period after diagnosis, possibly indicating longer potential survival. The algorithm needs to exemplify true short-term survivors, which would include individuals who indisputably passed away after a brief observation time. Consequently, when constructing the training dataset, it is necessary to exclude all individuals with a short survival period who are still alive.

To address this, we will omit the lower quartile of intervals for patients who were alive at their last observation (Fig. 1). This corresponds to 500 days for Overall Survival (OS). This approach aligns with the biological rationale: if a patient survives the initial year and a half, they can be utilized as a “long-liver” for training purposes. Similarly, for disease-free survival, the equivalent cutoff is the first 365 days from the initiation of therapy, as early relapses occur within the initial year. This methodology thus aligns with clinical significance.

**Fig. 1.**
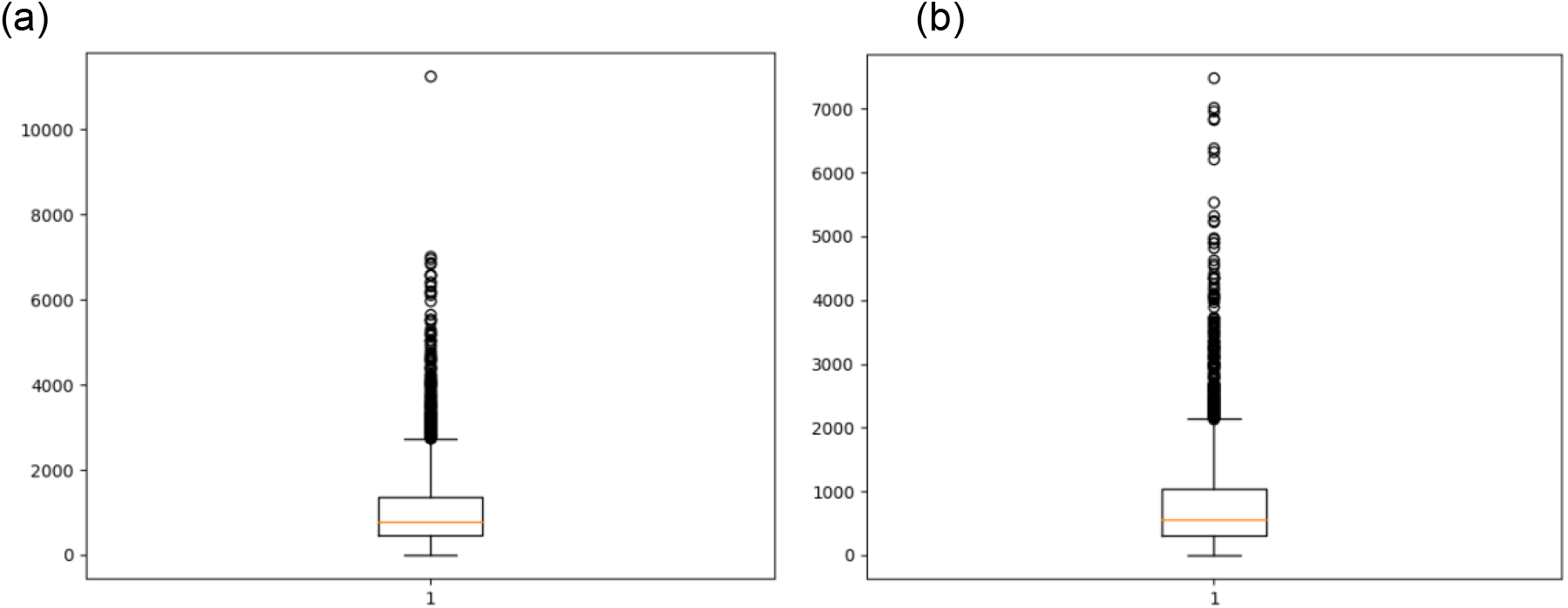
Patient datasets sorted by overall survival (a) and progression free interval (b)

**Fig. 2.**
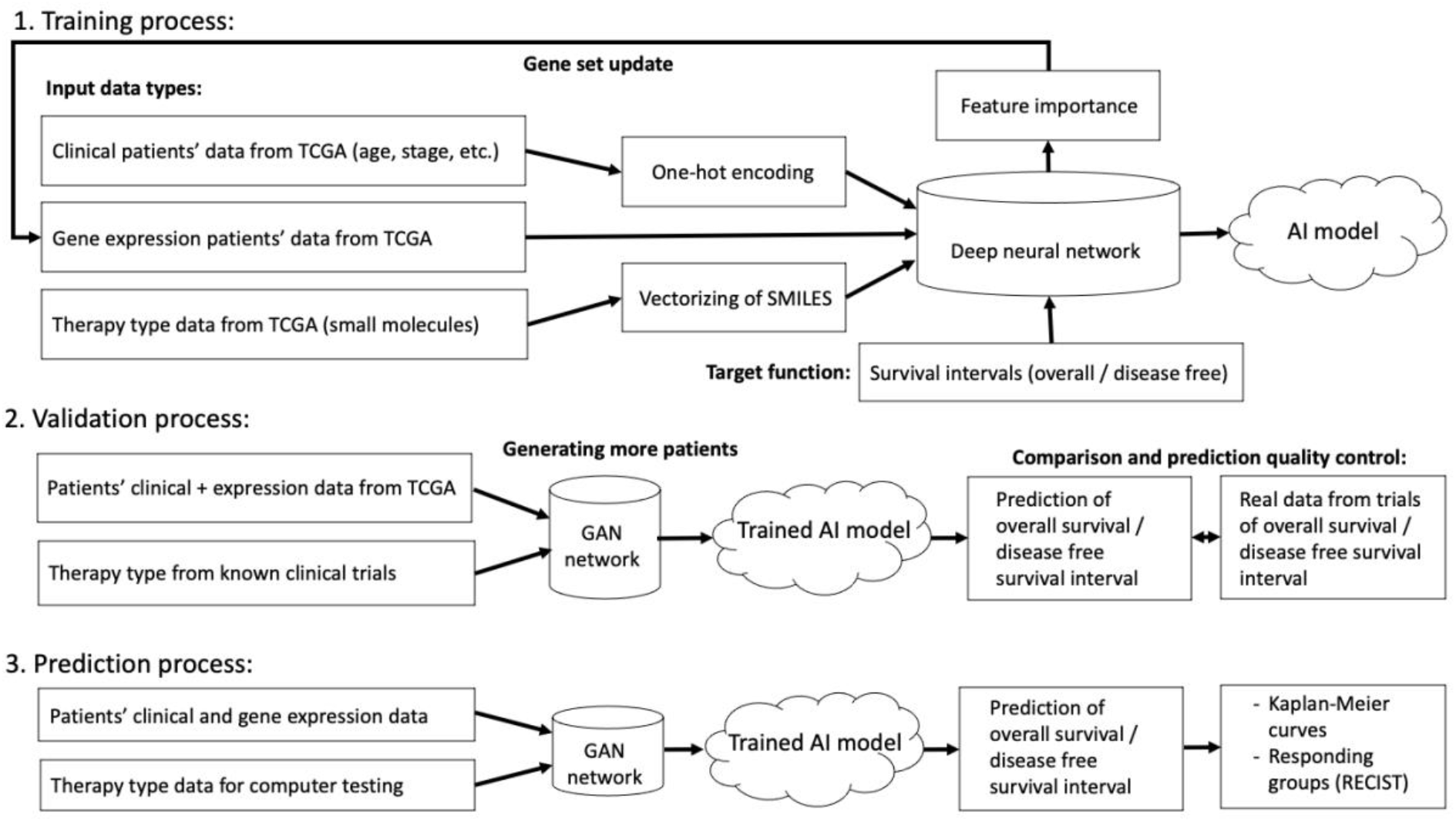
Overall workflow diagram

The gene expression data included no fewer than 20,000 genes. However, including all of them in the dataset would result in an imbalanced training set. For optimal training, the gene expression data should be reduced to several hundred genes.

To reduce dimensionality, we performed a feature importance analysis on the complete set of genes to select the top 150 most significant genes. We utilized the SHAP library for Python [16, 17] to determine the feature importances.

For the model we used the Keras library for deep learning. We trained our models to predict the number of months for overall survival (OS) and progression free interval (PFI).

We addressed three types of problems to solve:

1. Regression: Predicting the duration in months for OS and PFI.
2. Binary classification: Predicting the occurrence of events (death or recurrence) for OS and PFI.
3. Multi-class classification: Predicting the response type according to the RECIST scale (‘Complete’, ‘Partial’, ‘Progressive’, ‘Stable’).

For the prediction quality control we use 5-fold cross-validation.

It is necessary to compare our results with data from real clinical trials from open sources containing sufficient data for Kaplan-Meiers curve construction for OS and PFI [18, 19]. Let’s predict the outcomes of these trials by specifying into our approach their initial parameters: the diagnosis of the tested patients and the molecule(s) or their combination involved. Additionally, we extracted response categories to therapy from the clinicaltrials.gov.

As patient cohorts, we will assemble the necessary number of patients with known gene expression from the TCGA database. However, for certain diagnoses, the database contains a limited number of patients, which would hinder the construction of robust survival curves. To avoid this, we obtained additional synthetic patients’ data using generative adversarial networks (GAN) technology, which has been successfully used in various industries, such as image generation [20]. To implement GAN, we used the sdv library [21], in particular, the CTGAN module [22]. Thanks to this approach, we will generate the necessary amount of patient tabular data containing gene expression and clinical information.

## Results

1. Regression accuracy for OS and PFI intervals prediction after cross-validation is presented in Table 1.
2. Binary classification for predicting the occurrence of events (death or recurrence): OS - 0.77 ROC AUC, PFI - 0.73
3. Results of multi-class classification of the response type according to the RECIST scale are presented in Table 2.

To model real clinical trials with known outcomes, trials involving biological molecules were excluded from the available data, retaining only trials involving small molecules. Trials for three diagnoses were selected: breast cancer, lung cancer, and prostate adenocarcinoma. A total of 80 patient groups were utilized, including:

- Lung cancer: 28 trials, 38 pairs of curves
- Breast cancer (BC): 15 trials, 22 pairs of curves
- Prostate cancer: 6 trials, 11 pairs of curves

**Table 1.** Regression accuracy.

|  | Overall survival (OS) | Progression free interval (PFI) |
| --- | --- | --- |
| Pearsons correlation | 0.975 | 0.986 |
| R2 | 0.89 | 0.94 |
| RMSE | 6.05 | 4.52 |
| MAE | 3.85 | 3.71 |

**Table 2.** Multi-class classification accuracy.

|  | precision | recall | f1-score | Total patients |
| --- | --- | --- | --- | --- |
| Complete | 1.00 | 0.77 | 0.87 | 811 |
| Partial | 0 | 0 | 0 | 133 |
| Progressive | 0.60 | 0.56 | 0.58 | 435 |
| Stable | 0.54 | 1.00 | 0.70 | 223 |
| accuracy | 0.68 |  |  | 1602 |
| weighted average | 0.74 | 0.68 | 0.69 | 1602 |

In 8 cases, prediction failures occurred, resulting in the inability to construct curves. Figure 3 displays curves for real and predicted patient groups using the described approach. Figure 4 illustrates the distribution of Pearson correlations between real and predicted curves.

**Fig. 3.**
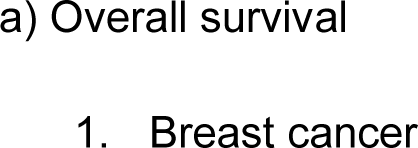
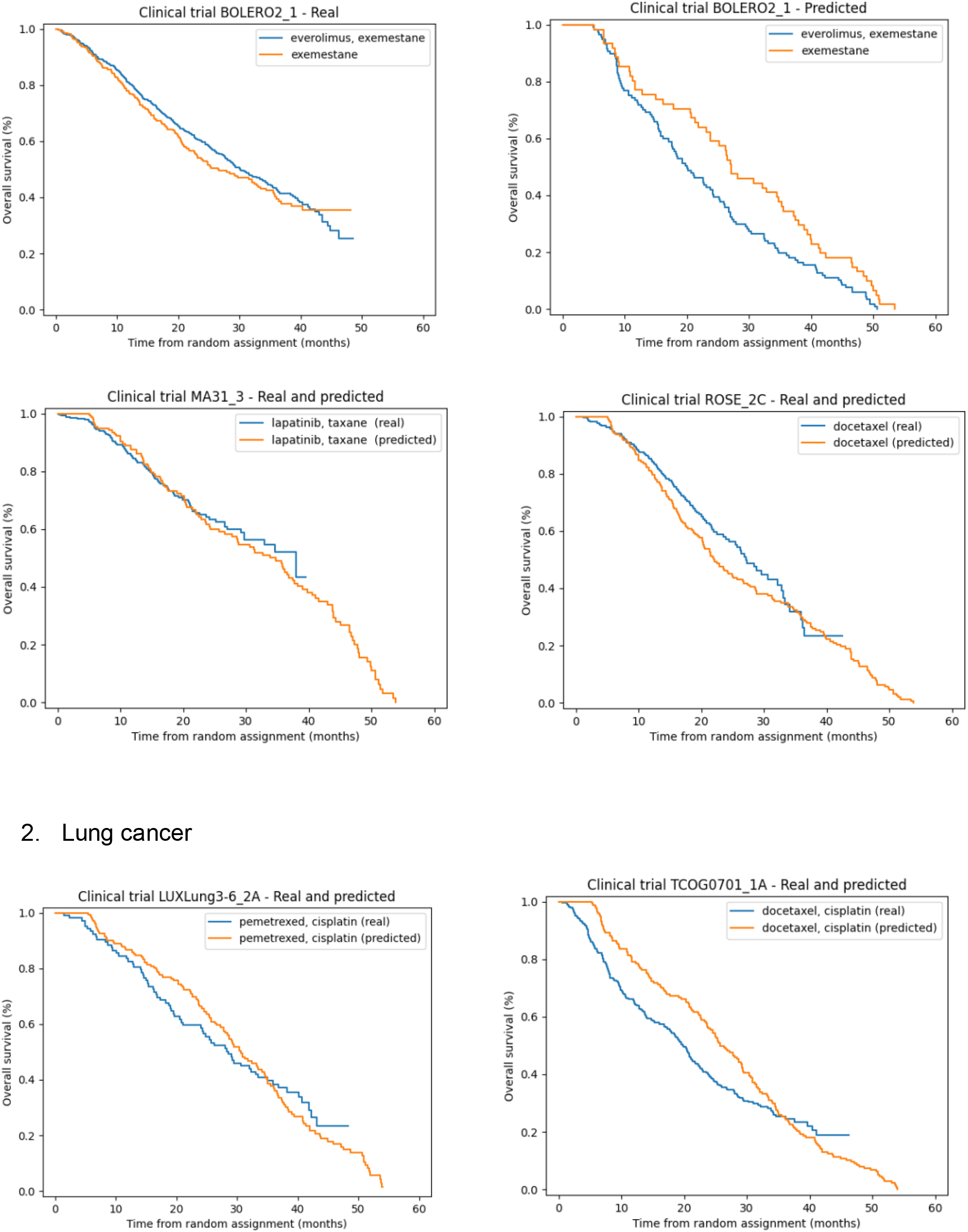
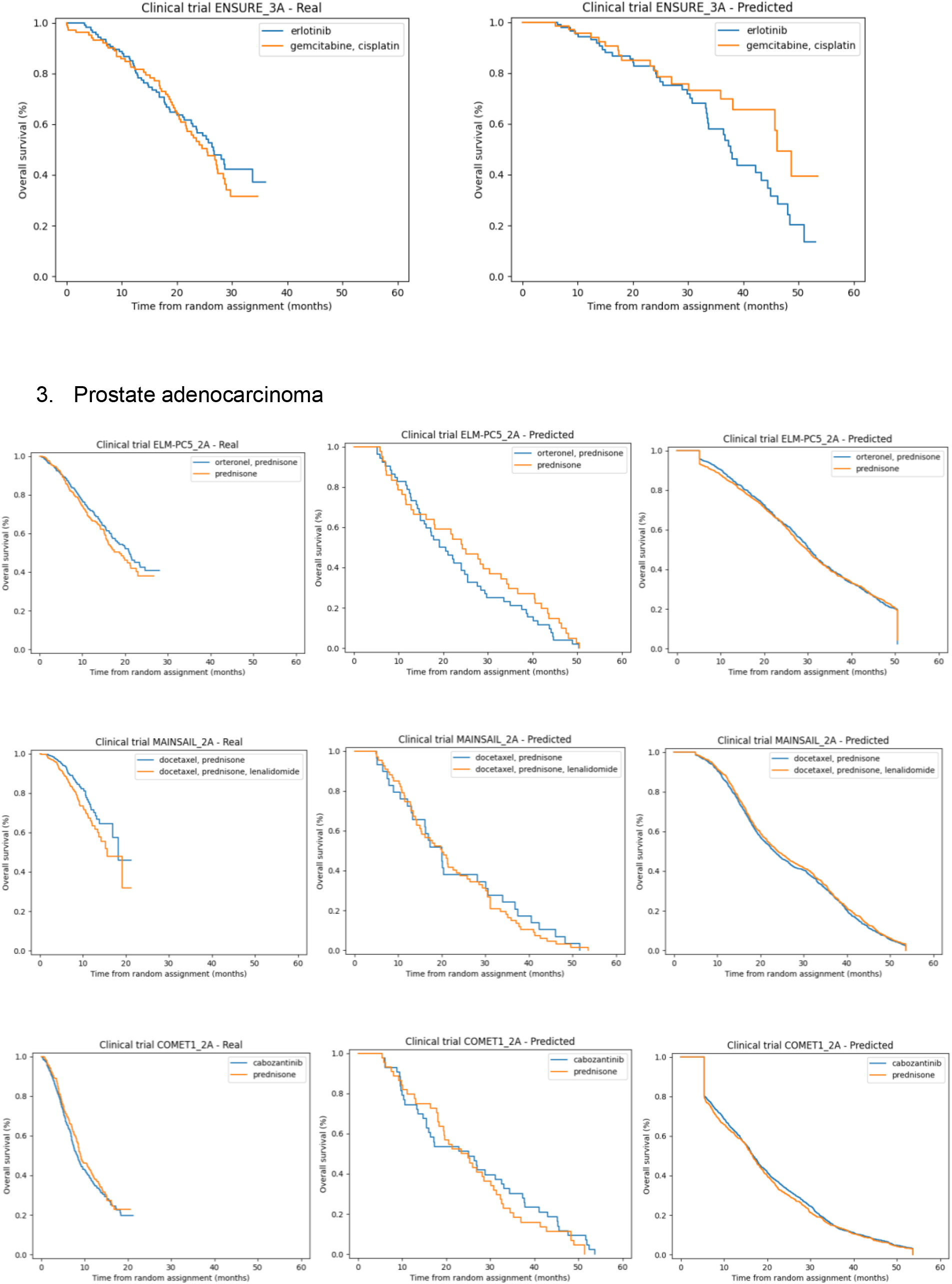
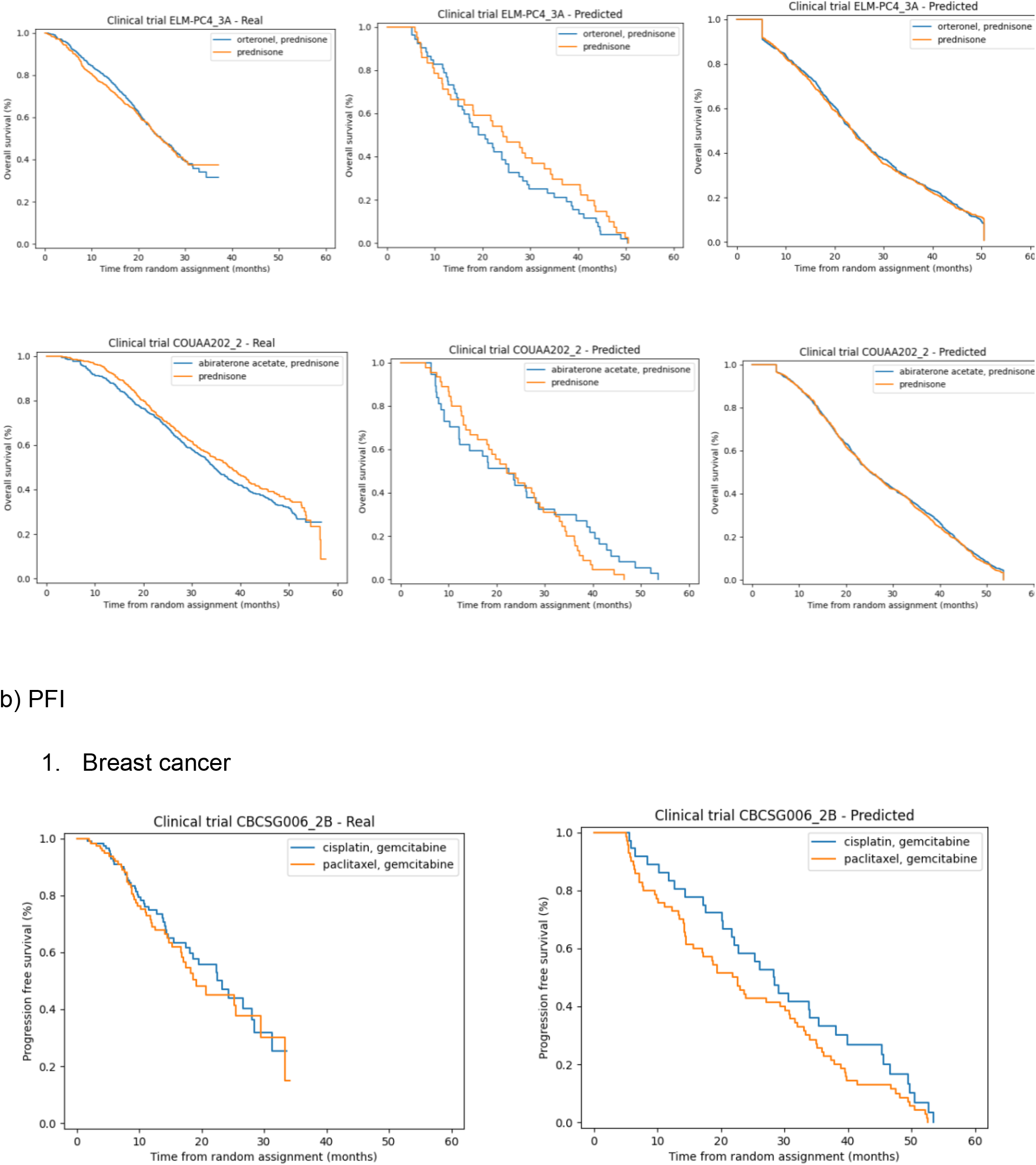
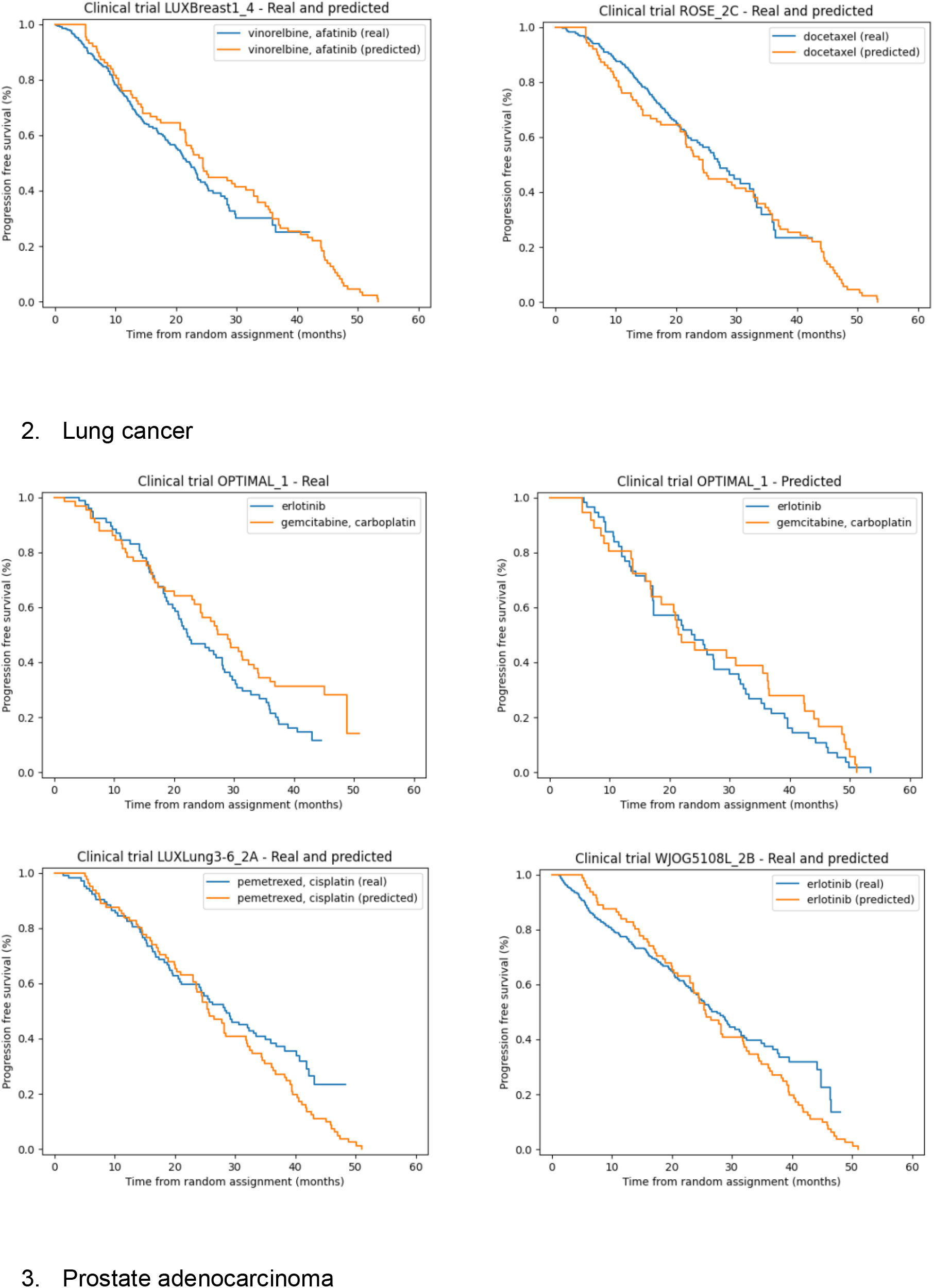
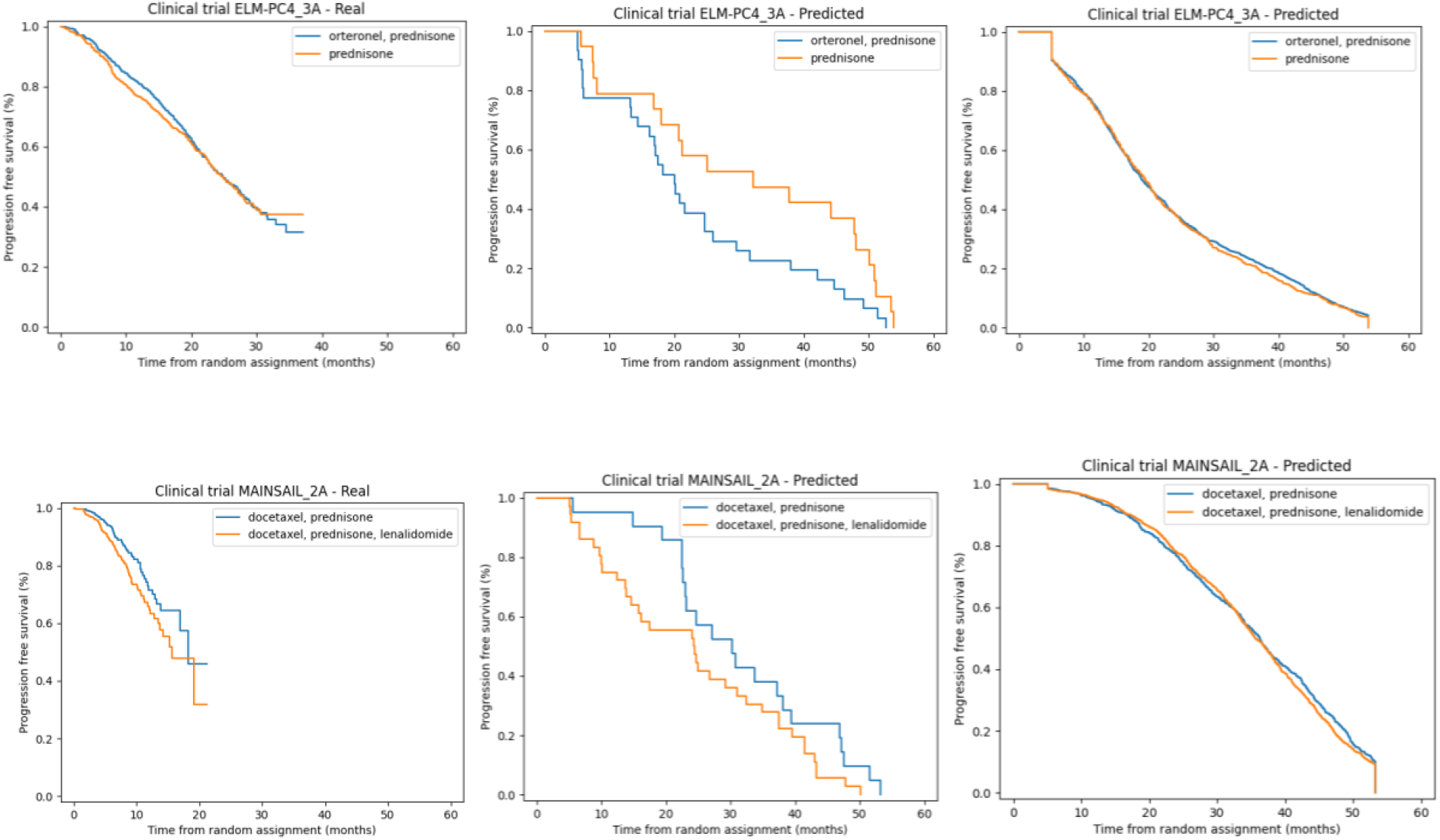
Kaplan-Meier curves for real and predicted clinical trials.

**Fig. 4.**
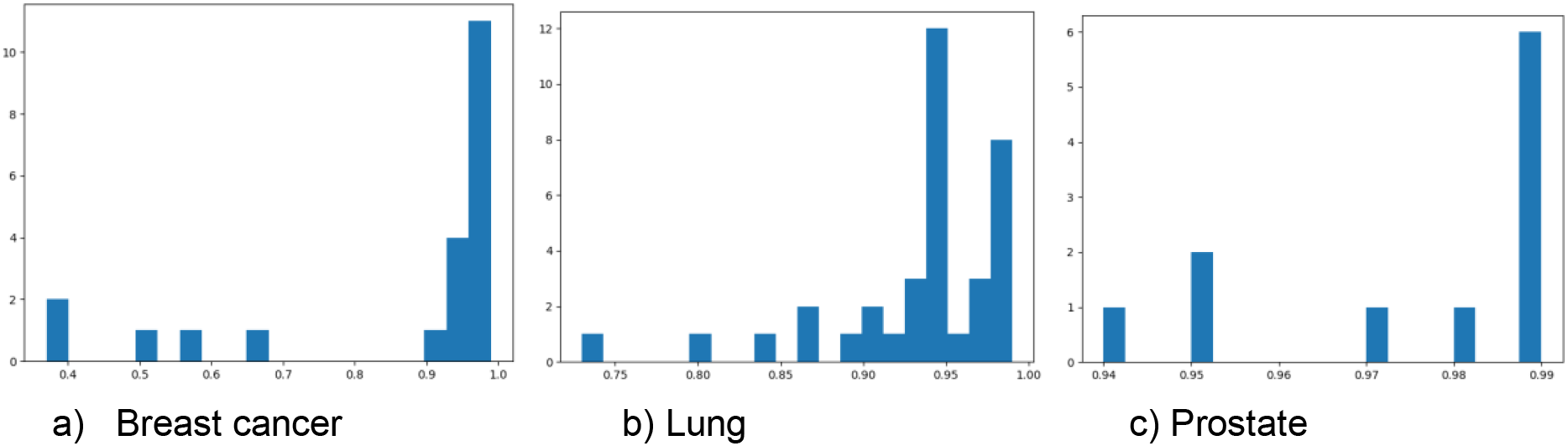
Pearson correlations between real and predicted Kaplan-Meier curves

Results of multi-class classification of the response type for the real clinical trials with known outcomes was conducted based on the percentage of objective responses (OR), as this metric was the only one available for the majority of trials. Consequently, the algorithm predicted four response categories on the RECIST scale mentioned above, from which the proportion of patients with OR was derived through summation. Prediction was performed for a total of 47 trials The following accuracy was achieved between these quantitative characteristics: mean average error (MAE) = 0.27 (95% CI: 0.21 - 0.33)

## Discussion

Utilization of vast and valuable datasets, such as patient OMICS data, remains a common challenge in clinical trial planning. Yet, the utilization of such data types allows for the consideration of molecular tumor characteristics, which, in turn, can aid researchers in identifying the necessary biomarkers as criteria for the application of newly developed drugs. While most new anticancer drugs possess biomarkers, they are primarily derived from preclinical studies and represent a fragmented mechanism targeted by the drug. However, such an approach fails to account for biomarker combinations, such as parallel oncogenic signaling pathways, potentially leading to resistance or recurrence due to selection based on a single marker.

The proposed approach aims to address this issue by identifying key features within patient cohorts, leveraging omics data available in the initial datasets, particularly gene expression. This also extends to synthetic patients generated by generative networks. At first glance, this approach may appear to generate non-existent patients, potentially distorting the trial modeling landscape. However, the generation of new patients is rooted in existing examples, resulting in the variances of attributes in the generated patients (e.g., gene expression, disease stage, or age) corresponding to the variances of these attributes in real patients. Consequently, with the expansion of the patient dataset, there is a high likelihood of the emergence of patients akin to those currently being artificially generated.

Visual analysis of Kaplan-Meier curve plots reveals that the method’s errors lie not in the quality of the plots but in quantitative characteristics. This signifies that, for more successful operation, a larger volume of data is required.

## Conclusion

The paper introduces a method for modeling clinical trials using omics data. It is designed to enhance the accuracy of protocol planning and interactively test hypotheses regarding the effectiveness and safety of newly developed drugs.

Based on the literature analysis, no approaches similar to the one presented have been found. This approach utilizes detailed patient-level data, full transcriptomic data, and incorporates clinical trial metrics as the objective function. The approach of generating synthetic patients has shown positive results, improving prediction accuracy both quantitatively and visually.

The presented approach, when implemented as a software platform, aims to expedite the drug discovery processes.

## Data Availability

All data produced in the present study are available upon reasonable request to the authors

## Notes

### Competing Interest Statement

Dmitrii K Chebanov, BioAlg Corp., shareholder

### Funding Statement

This study did not receive any funding

## References

1. Chebanov, D. K.; Tatevosova, N. S.; Mikhailova, I. N. Machine learning for predicting overall survival using whole exome DNA and gene expression data and analyzing the significance of features[abstract]. In: Proceedings of the AACR Virtual Special Conference on Artificial Intelligence, Diagnosis, and Imaging; 2021 Jan 13-14. Philadelphia (PA): AACR; Clin. Cancer. Res. 2021, 27(5_Suppl), Abstract nr PO-045

2. Borisov N, Sorokin M, Tkachev V, Garazha A, Buzdin A. Cancer gene expression profiles associated with clinical outcomes to chemotherapy treatments. BMC Med Genomics. 2020 Sep 18;13(Suppl 8):111. doi: 10.1186/s12920-020-00759-0. PMID: 32948183; PMCID: PMC7499993

3. Turki Turki, Jason T.L. Wang, Clinical intelligence: New machine learning techniques for predicting clinical drug response, Computers in Biology and Medicine, Volume 107, 2019, Pages 302–322, ISSN 0010-4825.

4. Vittrant B, Leclercq M, Martin-Magniette ML, Collins C, Bergeron A, Fradet Y, Droit A. Identification of a Transcriptomic Prognostic Signature by Machine Learning Using a Combination of Small Cohorts of Prostate Cancer. Front Genet. 2020 Nov 25;11:550894. doi: 10.3389/fgene.2020.550894. PMID: 33324443; PMCID: PMC7723980.

5. Tan AC, Naiman DQ, Xu L, Winslow RL, Geman D. Simple decision rules for classifying human cancers from gene expression profiles. Bioinformatics. 2005 Oct 15;21(20):3896–904. doi: 10.1093/bioinformatics/bti631. Epub 2005 Aug 16. PMID: 16105897; PMCID: PMC1987374.

6. Motwani M, Dey D, Berman DS, Germano G, Achenbach S, Al-Mallah MH, Andreini D, Budoff MJ, Cademartiri F, Callister TQ, Chang HJ, Chinnaiyan K, Chow BJ, Cury RC, Delago A, Gomez M, Gransar H, Hadamitzky M, Hausleiter J, Hindoyan N, Feuchtner G, Kaufmann PA, Kim YJ, Leipsic J, Lin FY, Maffei E, Marques H, Pontone G, Raff G, Rubinshtein R, Shaw LJ, Stehli J, Villines TC, Dunning A, Min JK, Slomka PJ. Machine learning for prediction of all-cause mortality in patients with suspected coronary artery disease: a 5-year multicentre prospective registry analysis. Eur Heart J. 2017 Feb 14;38(7):500–507. doi: 10.1093/eurheartj/ehw188. PMID: 27252451; PMCID: PMC5897836.

7. Subash Raj Susai, David Mongan, Colm Healy, Mary Cannon, Gerard Cagney, Kieran Wynne, Jonah F. Byrne, Connie Markulev, Miriam R. Schäfer, Maximus Berger, Nilufar Mossaheb, Monika Schlögelhofer, Stefan Smesny, Ian B. Hickie, Gregor E. Berger, Eric Y.H. Chen, Lieuwe de Haan, Dorien H. Nieman, Merete Nordentoft, Anita Riecher-Rössler, Swapna Verma, Rebekah Street, Andrew Thompson, Alison Ruth Yung, Barnaby Nelson, Patrick D. McGorry, Melanie Föcking, G. Paul Amminger, David Cotter, Machine learning based prediction and the influence of complement – Coagulation pathway proteins on clinical outcome: Results from the NEURAPRO trial, Brain, Behavior, and Immunity, Volume 103, 2022, Pages 50-60, ISSN 0889-1591

8. Chen W, Zhou C, Yan Z, Chen H, Lin K, Zheng Z, Xu W. Using machine learning techniques predicts prognosis of patients with Ewing sarcoma. J Orthop Res. 2021 Nov;39(11):2519–2527. doi: 10.1002/jor.24991. Epub 2021 Jan 24. PMID: 33458857.

9. Sherazi SWA, Jeong YJ, Jae MH, Bae JW, Lee JY. A machine learning-based 1-year mortality prediction model after hospital discharge for clinical patients with acute coronary syndrome. Health Informatics J. 2020 Jun;26(2):1289–1304. doi: 10.1177/1460458219871780. Epub 2019 Sep 30. PMID: 31566458.

10. Aliper, A., Kudrin, R., Polykovskiy, D., Kamya, P., Tutubalina, E., Chen, S., Ren, F. and Zhavoronkov, A. (2023), Prediction of Clinical Trials Outcomes Based on Target Choice and Clinical Trial Design with Multi-Modal Artificial Intelligence. Clin Pharmacol Ther.

11. Cancer Genome Atlas Research Network, Weinstein, J. N.; Collisson, E. A.; Mills, G. B.; Shaw, K. R.; Ozenberger, B. A.; Ellrott, K.; Shmulevich, I.; Sander, C.; Stuart, J. M. The cancer genome atlas pan-cancer analysis project. Nat. Genet. 2013, 45(10), 1113–1120.

12. Wishart, D. S.; Feunang, Y. D.; Guo, A. C.; Lo, E. J.; Marcu, A.; Grant, J. R.; Sajed, T.; Johnson, D.; Li, C.; Sayeeda, Z.; Assempour, N.; Iynkkaran, I.; Liu, Y.; Maciejewski, A.; Gale, N.; Wilson, A.; Chin, L.; Cummings, R.; Le, D.; Pon, A.; Knox, C.; Wilson, M. DrugBank 5.0: a major update to the DrugBank database for 2018. Nucleic Acids Res. 2017, 8.

13. RDKit: Open-source cheminformatics; http://www.rdkit.org

14. Jaeger, S.; Fulle, S.; Turk, S. Mol2vec: Unsupervised Machine Learning Approach with Chemical Intuition?. J. Chem. Inf. Model. 2018, 58(1), 27–35.

15. Mikolov, T.; Chen, K.; Corrado, G.; Dean, J. Efficient Estimation of Word Representations in Vector Space. arXiv 2013, 1301.3781.

16. Lundberg, S.M., Nair, B., Vavilala, M.S. et al. Explainable machine-learning predictions for the prevention of hypoxaemia during surgery. Nat Biomed Eng 2, 749–760 (2018). 10.1038/s41551-018-0304-0

17. Lundberg, S. M, Lee, S.-I. A Unified Approach to Interpreting Model Predictions. Advances in Neural Information Processing Systems 30, 4765--4774 (2017), Curran Associates, Inc. http://papers.nips.cc/paper/7062-a-unified-approach-to-interpreting-model-predictions.pdf

18. Cancer patient survival can be parametrized to improve trial precision and reveal time-dependent therapeutic effects” Deborah Plana, Geoffrey Fell, Brian M. Alexander, Adam C. Palmer, Peter K. Sorger. Nat Commun 13, 873 (2022). 10.1038/s41467-022-28410-9

19. Plana, D., Fell, G., Alexander, B. M., Palmer, A. C. & Sorger, P. K. Imputed individual participant data from oncology clinical trials. Synpase repository. 10.7303/SYN25813713 (2021)

20. Goodfellow I. J.; Pouget-Abadie J.; Mirza M.; Xu B.; Warde-Farley, D.; Ozair, S.; Courville, A.; Bengio, Y. Generative adversarial networks. arXiv, 2014. 1406.2661.

21. Patki, N.; Wedge, R.; Veeramachaneni, K. The Synthetic Data Vault, 2016 IEEE International Conference on Data Science and Advanced Analytics (DSAA) 2016, 399–410.

22. Xu, L.; Skoularidou, M.; Cuesta-Infante, A.; Veeramachaneni, K. Modeling Tabular data using Conditional GAN. NeurIPS, 2019.

